# Measuring Repositioning in Home Care for Pressure Injury Prevention and Management

**DOI:** 10.1101/2022.03.18.22272369

**Authors:** Sharon Gabison, Nikola Pupic, Gary Evans, Elham Dolatabadi, Geoff Fernie, Tilak Dutta

## Abstract

A critical best practice for prevention and management of pressure injuries is regularly repositioning individuals who are at risk of these injuries are when they are in bed. However, despite the widespread agreement of the need for regular repositioning (typically every two hours), adherence to repositioning schedules remains poor in the clinical environment and there are some indications that the situation in home environment is even worse.

Our team has recently developed a non-contact system that can continuously determine an individual’s position in bed (left-side lying, right-side lying or supine) using data from a set of inexpensive load cells placed under the bed frame. A proof of principle study showed that our system was able to detect whether healthy participants were supine, left-side lying or right-side lying with 94.2% accuracy in the lab environment.

The objective of the present work was to deploy our system into the home environment to evaluate how well the system was able to detect the position of individuals sleeping in their own beds overnight by comparing to ground truth time-lapse camera images using eight machine learning classifiers. Nine participants were recruited for this study and we found our system was able to detect an individual’s position in bed with 98.1% accuracy and an F1 score of 0.982 using the XGBoost classifier.

Future work will include using this system to evaluate interventions focused on improving adherence to 2-hour repositioning schedules for pressure injury prevention or management as well as incorporating this technology in a repositioning prompting system to alert caregiver when a patient has remained in the same position for too long.

## 1 INTRODUCTION

Pressure injuries are caused by damage to the skin and underlying soft tissue due to prolonged tissue deformation [1] and can cause septicemia leading to death in the most severe cases. Pressure injuries can be a devastating secondary complication of immobility [2]. These injuries cause pain and suffering to those who live with them and increase caregiver burden [3]. The costs associated with pressure injuries are staggering; annual costs of hospital acquired pressure injuries in the United States are estimated to exceed $26.8 billion [4]. It is not surprising that the costs of preventing a pressure injury are much lower than the costs to manage them [5].

Many, but not all pressure injuries are preventable [6]. A key best practice guideline for prevention and management of pressure injuries is to reposition at-risk patients every two hours to enable compressed tissues to return to their normal shape [7]. In the case of individuals being unable to reposition themselves, they often rely on their caregiver to assist them. However, adherence to pressure offloading schedules has been found to be poor in the clinical environment, and may be worse in the home care environment where there is evidence that awareness of these injuries and the importance of repositioning is lacking [8]. There is a strong need for the development of sensor systems that can detect how often patients are being repositioned so that interventions to improve repositioning frequency can be evaluated objectively.

There have been a number of recent advanced using machine learning for predicting an individual’s position and/or movements in bed [9-11]. These methods include load cells placed under the legs of a bed [10, 12, 13], pressure mats positioned over the mattress [11, 14], temperature sensors [15], automated image analysis systems [9], single motion inertial sensor units [16] and capacitive ECG signals [17]. However, there are limitations to each of these systems and the context in which they have been evaluated. For example, automated image analysis may be difficult to predict an individual’s position in bed if there is occlusion of the individual by a blanket. Furthermore, with image analysis, there is a the need to train classifiers for each position in bed, making the detection of arbitrary positions challenging [9]. Methods that rely on single use inertial motion units require that the sensors attach to an individual’s chest using adhesive which can potentially lead to skin damage in an individual with compromised skin integrity. Many of these methods have also been tested in controlled laboratory environments using predetermined positions [10] [12] [13] and do not incorporate the wide range of natural sleep positions adopted in bed, resulting in limited generalizability of findings to the natural environment. Furthermore, existing studies have largely not included participants who are at risk of developing a pressure injury.

Our team has developed a patient position detection system that is designed to detect whether an individual in bed is on their left side, right side or supine. This system uses data from four load cells placed under the legs of the bed. Sensors collect force data and features are extracted from the data. Using advanced machine learning algorithms, we have been able to predict an individual’s position in bed as either supine, right-side-lying or left-side-lying with 94.2% accuracy in an controlled laboratory environment [18]. Our ultimate goal is to build on this position detection system to develop a repositioning prompting system that will continuously monitor an patient’s position in bed and alert a caregiver when the patient has remained in the same position for too long (e.g. 2 hours) so they an be repositioned regularly.

To-date, our system has not been tested in the home environment, where individuals sleep naturally in their own bed. The primary objective of this study was to evaluate our system’s ability to predict whether participants were on their left side, right side or supine while the individual slept naturally in their own bed. The secondary objective was to measure the inter-rater reliability of labelling ground truth participant position from visual inspection of time-lapse images captured of the individuals sleeping.

## 2 METHODS

This study was approved by the Research Ethics Board of University Health Network.

### Participants

A convenience sample of 9 healthy participants (4 males, 5 females) were recruited through advertisements placed throughout Toronto Rehabilitation Institute – University Health Network. Inclusion criteria included access to a single bed at home supported from four points below the bed. Interested participants contacted the study team via email or phone. Once participants were deemed eligible for the study and consented to participate, an overnight data collection session was scheduled.

### System Setup and Data Collection

Data was collected from participants while they slept overnight in their single bed at home. Four single axis load cells (each comprised of four strain gauges, model DLC902-30KG-HB, resolution of 1.8 g with an error of 0.2%, Hunan Detail Sensing Technology, Changsha, Hunan, China, arranged to create a full Wheatstone bridge) were placed under the legs of the bed. Signals from the load cells were amplified (gain of 500), filtered and converted from analog to digital values (5.0 VDC excitation) using a signal conditioner (GEN 5, AMTI, Watertown, MA). NetForce software (version 3.5.2, AMTI, Watertown, MA) was used on a PC Laptop (Thinkpad T520, Lenovo, Hong Kong, China, 2.5 GHz Intel Core i5 CPU, 4 GB RAM, 16 bit resolution) to collect and filter the data at 50 Hz. An infrared time-lapse camera mounted on a tripod at the foot of the bed captured an image of the participants every 30 seconds for ground truth labelling.

### 2.1 Data Processing

Vertical forces from each of the four load cells (LH: left sensor at the head of the bed, RH: right sensor at the head of the bed, LF: left sensor at the foot of the bed, RF: right sensor at the foot of the bed) were extracted and used with the distance between the sensors (length, *l*, and width, *w*) to compute the 12 features identified in Table 1.

**Table 1.** Features calculated from load cell data

| Feature | Description |
| --- | --- |
| mean CoM<sub>x</sub> | The mean of CoM<sub>x</sub> parallel to the width of the bed |
| meanCoM<sub>y</sub> | The mean of CoM<sub>y</sub> parallel to the length of the bed |
| ratio_meanCoM | The quotient of meanCoM<sub>y</sub> divided by CoM<sub>x</sub> |
| stdCoM<sub>x</sub> | The standard deviation of CoM<sub>x</sub> |
| stdCoM<sub>y</sub> | The standard deviation of CoM<sub>y</sub> |
| ratio_stdCoM | The quotient of stdCoM<sub>y</sub> divided by stdCoM<sub>x</sub> |
| CoM_resp_ANG | CoM angle during inhalation phase only, averaged for all occurrences |
| stdCoM_resp_ANG | Standard deviation of CoM_resp_ANG |
| rmsCoM_resp_x | The root mean square of the x-component of CoM_resp during both inhale and exhale phases, normalized to the 97<sup>th</sup> percentile |
| rmsCoM_resp_y | The root mean square of the y-component of CoM_resp during both inhale and exhale phases, normalized to the 97<sup>th</sup> percentile |
| ratio_rmsCoM_resp | The quotient of rmsCoM_resp_y divided by rmsCoM_resp_x |
| rmsPulse | The root mean square of the load cell signals filtered to capture changes from the cardiac cycle |

The two-dimensional centre of mass (CoM) of the participant-bed system was calculated using equations 1 and 2.

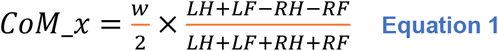

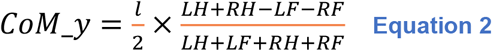

The CoM signal was lowpass filtered using a Chebyshev Type II filter to isolate the respiratory signal. The filter was applied using the filtfilt function that ensures a zero-phase shift to obtain the CoM_resp_x and CoM_resp_y. The times when maxima (tmax) and minima (tmin) occurred in the CoM_resp_x and CoM_resp_y signals were found by finding zero crossings for the first derivative of each signal. These times correspond with the end of each exhalation and inhalation respectively [54]. The angle of the principal axis of the ellipsoid traced by the resultant CoM_resp signal relative to the positive x axis (positive angle measured clockwise) was calculated using Equation 3 for each tmax and subsequent tmin.

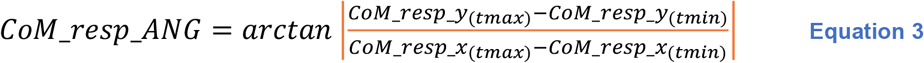

The changes from the components of the cardiac signal were isolated using an Equiripple FIR filter. The filtfilt function was used to bandpass filter the sum of the LH and RH signals ensuring a zero-phase shift.

Our data was processed similar to Wong et al. [18]. Each data point used for training/testing our machine learning classifiers was the average of a 45 s moving window that contained 2250 observations, where a new value was computed by shifting the window by 15 s.

#### Ground Truth Participant Position Labels

Three members of the research team independently reviewed the time-lapse images for each data point and assigned a label indicating the participant was in one of three positions: right-side-lying, left-side-lying, or supine for each 45 s. The most common label from the three reviewers was selected as the final ground truth label.

### 2.2 Data Analyses

#### System Accuracy

A leave-one-participant-out cross validation approach was used to evaluating the F1 score and accuracy of all classifiers. Using this approach, the classifiers were trained on data from 8 participants and tested on the remaining held-out participant. The procedure was repeated 9 times (once for each participant). This validation approach was selected to maximize the number of training observations that were used for each classifier.

#### Incremental Learning

An incremental learning approach was used to evaluate the ability of the classifier to adapt to the data from the left-out participant in a similar manner as described in Wong et al. [18]. The machine learning classifier was iteratively trained using different percentages of the left-out participant’s data (c, with c = 0%, 10%, 20%, or 30%). The left-out participant data was split into an incremental learning set and a test set with a ratio of 30:70 to ensure the test set was the same across all incremental learning levels (c values) and that the performances could be compared. For example, a *c* = 20%, reflects 20% of the overall left-out participant’s data being added into the training set to customize the model for that specific participant. This 20% of the left-out participant’s data is found in the incremental learning set that holds 30% of the overall left-out participant’s data.

#### Machine Learning Approach

The following 8 machine learning classifiers were used for analysis: AdaBoost (ADA), Gradient Boosting (GBC), Light Gradient Boosting (LGB), Logistic Regression (LR), two Multilayer Perceptrons (MLP), Support Vector Machine (SVM), and XGBoost (XGB). The first MLP model, MLP 1, was the model used by Wong et al. [18] and it is depicted in Figure 1. Table 2 depicts the structure of the model in more detail. The simplified MLP model, MLP 2, is depicted in Figure 2. Table 3 depicts the structure of the final simplified MLP model in more detail.

**Table 2.**
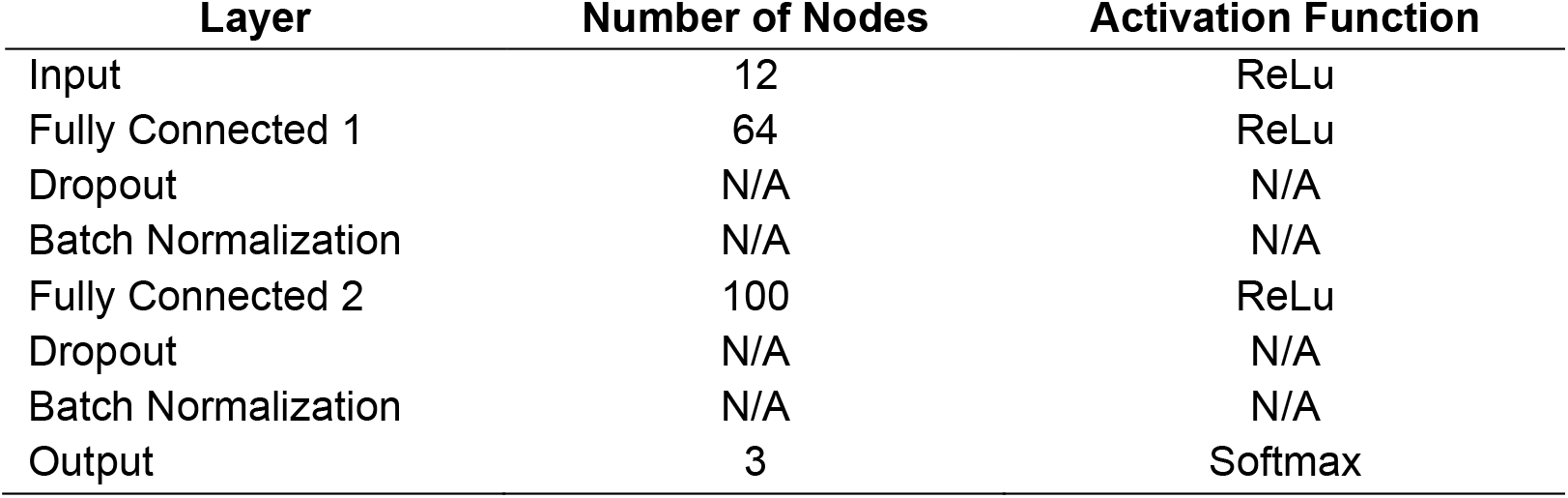
Architecture of the original MLP model by Wong et al.

| Layer | Number of Nodes | Activation Function |
| --- | --- | --- |
| Input | 12 | ReLu |
| Fully Connected 1 | 64 | ReLu |
| Dropout | N/A | N/A |
| Batch Normalization | N/A | N/A |
| Fully Connected 2 | 100 | ReLu |
| Dropout | N/A | N/A |
| Batch Normalization | N/A | N/A |
| Output | 3 | Softmax |

**Table 3.**
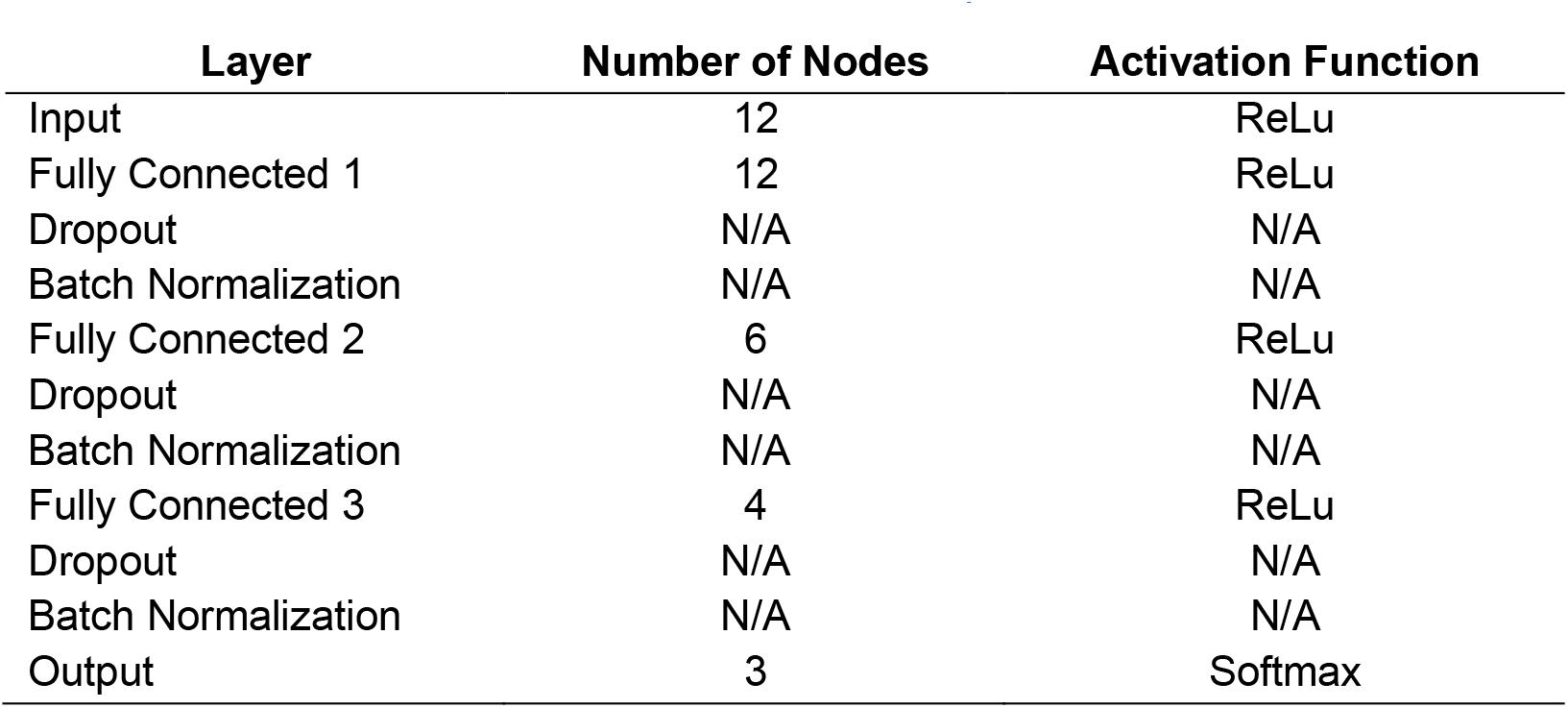
Architecture of the final simplified MLP model.

**Figure 1.**
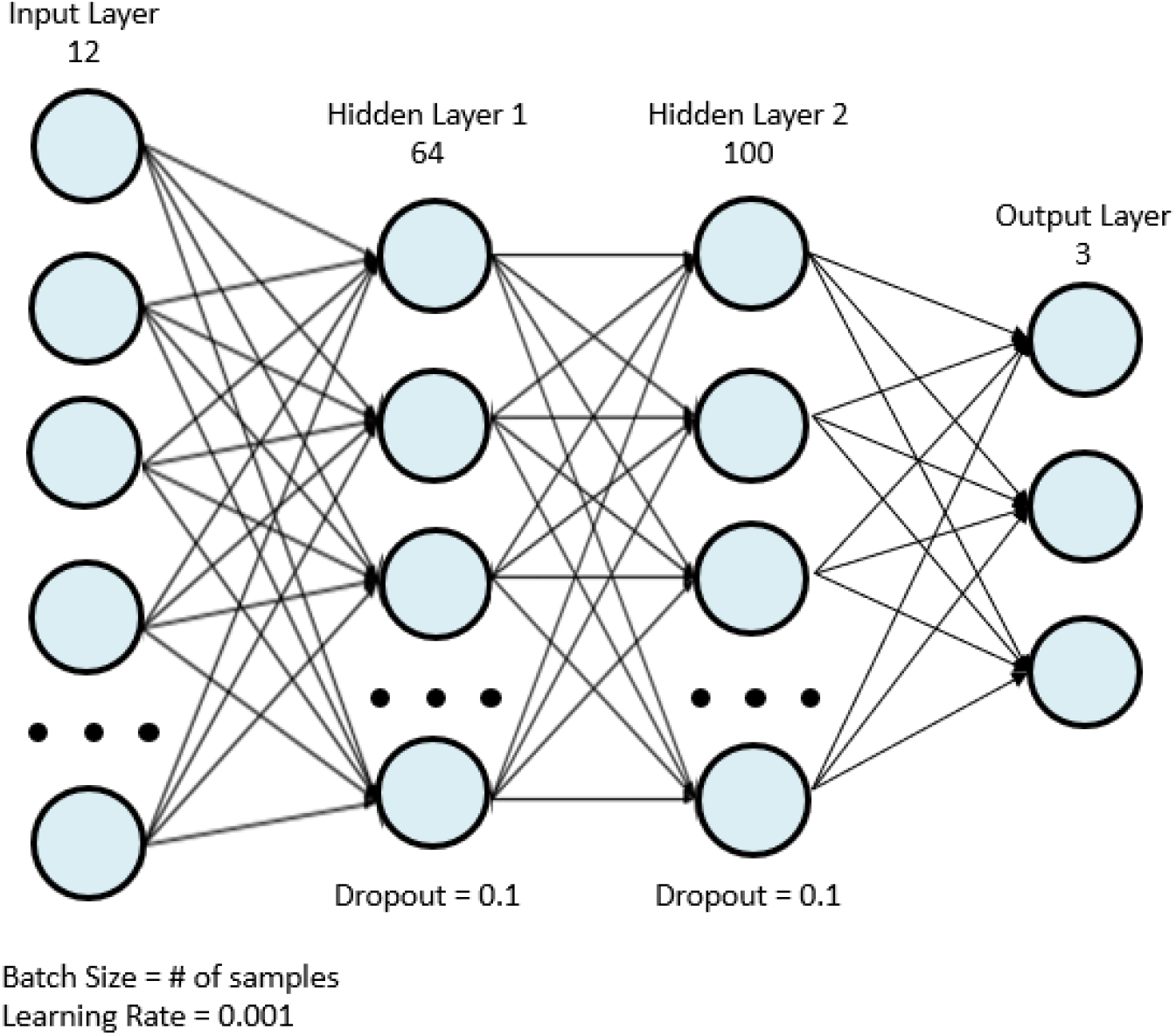
Original MLP used in the in-lab study by Wong et al.

**Figure 2.**
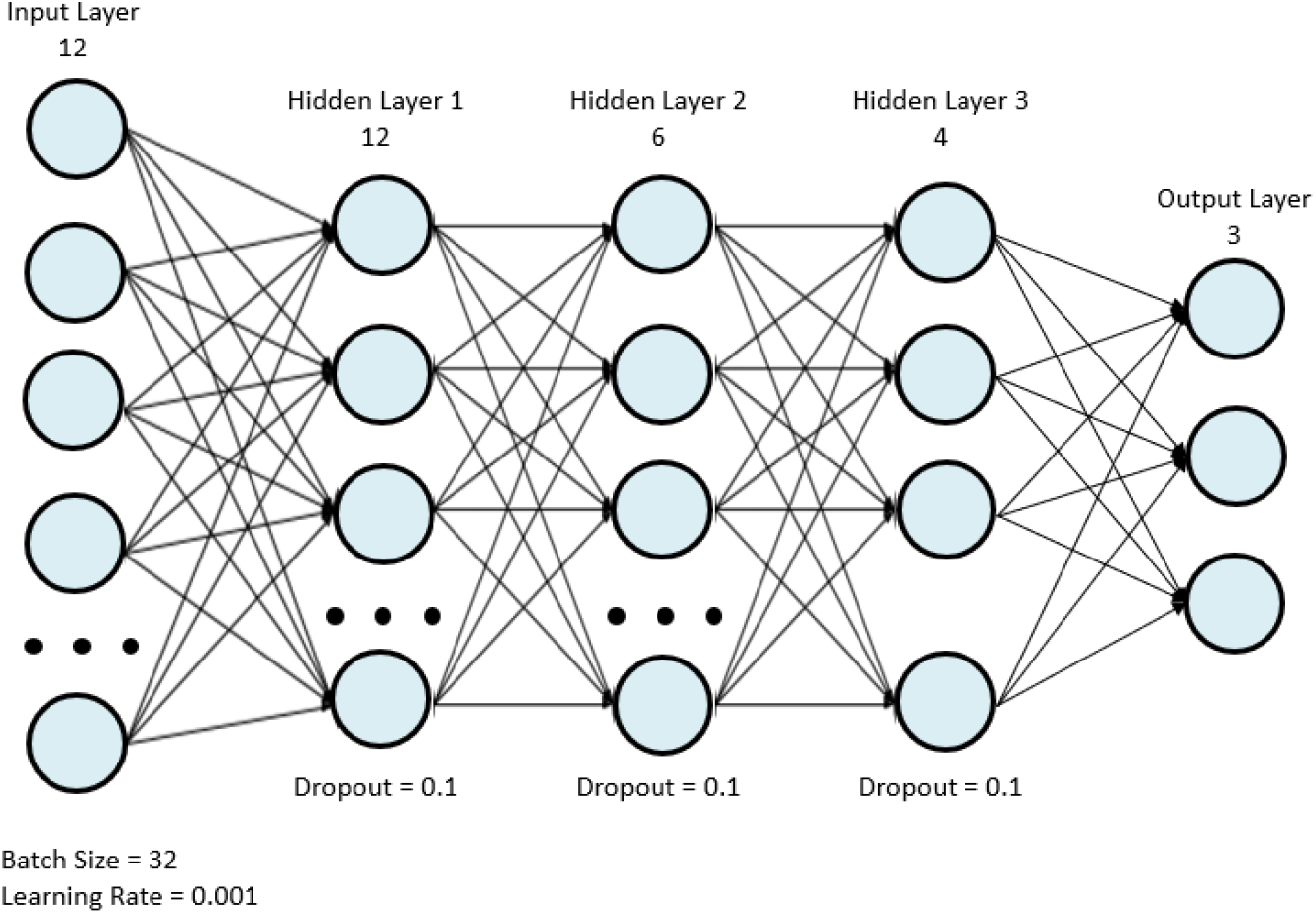
Final simplified MLP model.

### 2.3 Statistical Analyses

The performance of machine learning classifiers was analyzed using R Studio (R Studio, PBC, USA) and inter-rater reliability was analyzed using SPSS Statistics (SPSS Statistics 23, IBM, USA).

#### Machine Learning Classifiers

A Shapiro-Wilk test and a Levene test were used to assess for normality and homogeneity of the mean accuracies and F1 scores of the different models, respectively. A Friedman’s ANOVA was used to determine if there were significant differences between the mean accuracies and F1 scores of the machine learning classifiers at the *c* = 30% incremental learning. Post-hoc Wilcoxon Signed-Rank tests with Bonferroni adjustments were used to compare the top three performing models. For the top two performing models, the incremental learning levels corresponding to *c* = 0%, 10%, 20%, and 30% were compared using a Friedman’s ANOVA to determine how the level of incremental learning affected performance. Post-hoc Wilcoxon Signed-Rank tests with Bonferroni adjustments were used to compare the performance of the models to their adjacent incremental learning value(s) (i.e., 0% to 10%, 10% to 20%, 20% to 30%).

#### Inter-Rater Reliability

In order to analyze the reliability of ground truth data labelling of the three raters, an inter-rater reliability analysis was conducted using Fleiss Multirater Kappa statistics for each position, for each participant and for all combined participants and positions. The following criteria was used for reliability estimates: kappa < 0.20 representing poor agreement, kappa = 0.21 – 0.40 representing fair agreement, kappa = 0.41 – 0.60 representing moderate agreement, kappa = 0.61 – 0.80 representing good agreement, kappa = 0.81 – 1.00 representing almost perfect agreement [19].

## 3 RESULTS

### 3.1 Participants

Descriptive statistics of our participants are shown in Table 4.

**Table 4.** Participant descriptive statistics

| Participant | Sex (M/F) | Age Range (years) | Height (cm) | Weight (kg) | BMI |
| --- | --- | --- | --- | --- | --- |
| 1 | M | 70-79 | 171.5 | 70.0 | 23.9 |
| 2 | F | 70-79 | 160.0 | 56.8 | 22.1 |
| 3 | F | 70-79 | 170.0 | 61.3 | 21.2 |
| 4 | M | 30-39 | 180.3 | 84.1 | 25.9 |
| 5 | F | 50-59 | 157.0 | 85.0 | 34.5 |
| 6 | M | 20-29 | 167.0 | 69.5 | 24.5 |
| 7 | F | 60-65 | 156.0 | 61.4 | 25.2 |
| 8 | F | 60-65 | 162.0 | 54.4 | 20.7 |
| 9 | M | 60-69 | 181.0 | 80.5 | 24.5 |

### 3.2 Machine Learning Models

The overall mean accuracy and F1 scores and their standard deviation values across all nine participants for the classification of *left, right*, or *supine* at each incremental learning level for each model are represented in Tables 5 and 6. Figures 3 and 4 show the mean accuracy for each model at 30% incremental learning level. Further comparisons of performance were done at the 30% incremental learning level since these classifiers consistently outperformed the other incremental learning levels.

**Table 5.** Combined mean accuracy and standard deviations of the tested models for classifying positions as supine, left, or right.

| Model | c = 0% | c = 10% | c = 20% | c = 30% |
| --- | --- | --- | --- | --- |
| ADA | 62.27% ± 15.44% | 66.68% ± 12.21% | 71.86% ± 10.33% | 74.37% ± 11.09% |
| GBC | 62.27% ± 13.45% | 79.09% ± 8.66% | 84.98% ± 6.33% | 90.13% ± 4.64% |
| LGB | 61.68% ± 13.25% | 93.94% ± 4.52% | 96.86% ± 2.00% | 97.99% ± 1.42% |
| LR | 67.77% ± 16.01% | 69.05% ± 15.05% | 70.18% ± 14.34% | 71.23% ± 13.81% |
| MLP | 67.92% ± 14.95% | 69.35% ± 17.90% | 73.06% ± 15.29% | 75.40% ± 10.68% |
| OG | 64.11% ± 16.01% | 78.25% ± 10.91% | 85.69% ± 6.70% | 88.58% ± 6.27% |
| SVM | 71.25% ± 15.84% | 72.21% ± 15.54% | 73.09% ± 15.21% | 74.01% ± 14.85% |
| XGB | 61.75% ± 14.02% | 93.40% ± 5.12% | 96.83% ± 1.82% | 98.12% ± 1.03% |

**Table 6.** Combined F1 scores ± standard deviations of the tested models for classifying participant positions as supine, left, or right.

| Model | c = 0% | c = 10% | c = 20% | c = 30% |
| --- | --- | --- | --- | --- |
| ADA | 0.654 ± 0.153 | 0.697 ± 0.1240 | 0.7436 ± 0.1100 | 0.766 ± 0.1190 |
| GBC | 0.650 ± 0.147 | 0.808 ± 0.0813 | 0.8637 ± 0.0578 | 0.910 ± 0.0369 |
| LGB | 0.639 ± 0.138 | 0.946 ± 0.0361 | 0.9713 ± 0.0152 | 0.981 ± 0.0116 |
| LR | 0.695 ± 0.168 | 0.710 ± 0.1540 | 0.7225 ± 0.1440 | 0.733 ± 0.1370 |
| MLP | 0.695 ± 0.153 | 0.708 ± 0.1880 | 0.7441 ± 0.1500 | 0.769 ± 0.1120 |
| OG | 0.662 ± 0.161 | 0.802 ± 0.0901 | 0.8728 ± 0.0494 | 0.898 ± 0.0470 |
| SVM | 0.713 ± 0.167 | 0.723 ± 0.1640 | 0.7317 ± 0.1590 | 0.741 ± 0.1540 |
| XGB | 0.637 ± 0.154 | 0.941 ± 0.0403 | 0.9707 ± 0.0144 | 0.982 ± 0.0030 |

**Figure 3.**
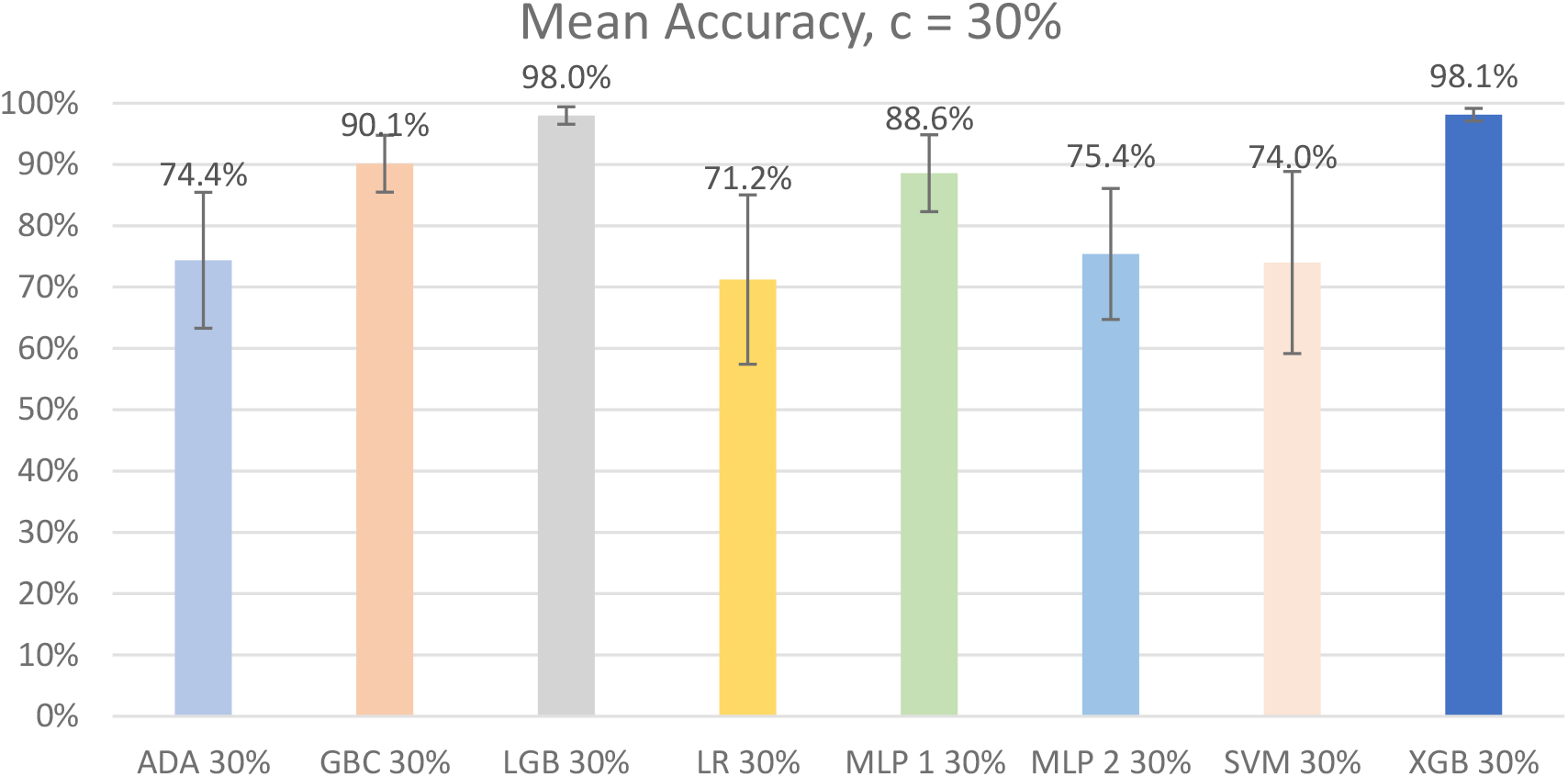
Graph of mean accuracy of Left, Right, or Supine classification by model for the incremental learning level c = 30%. Error bars represent standard deviations.

**Figure 4.**
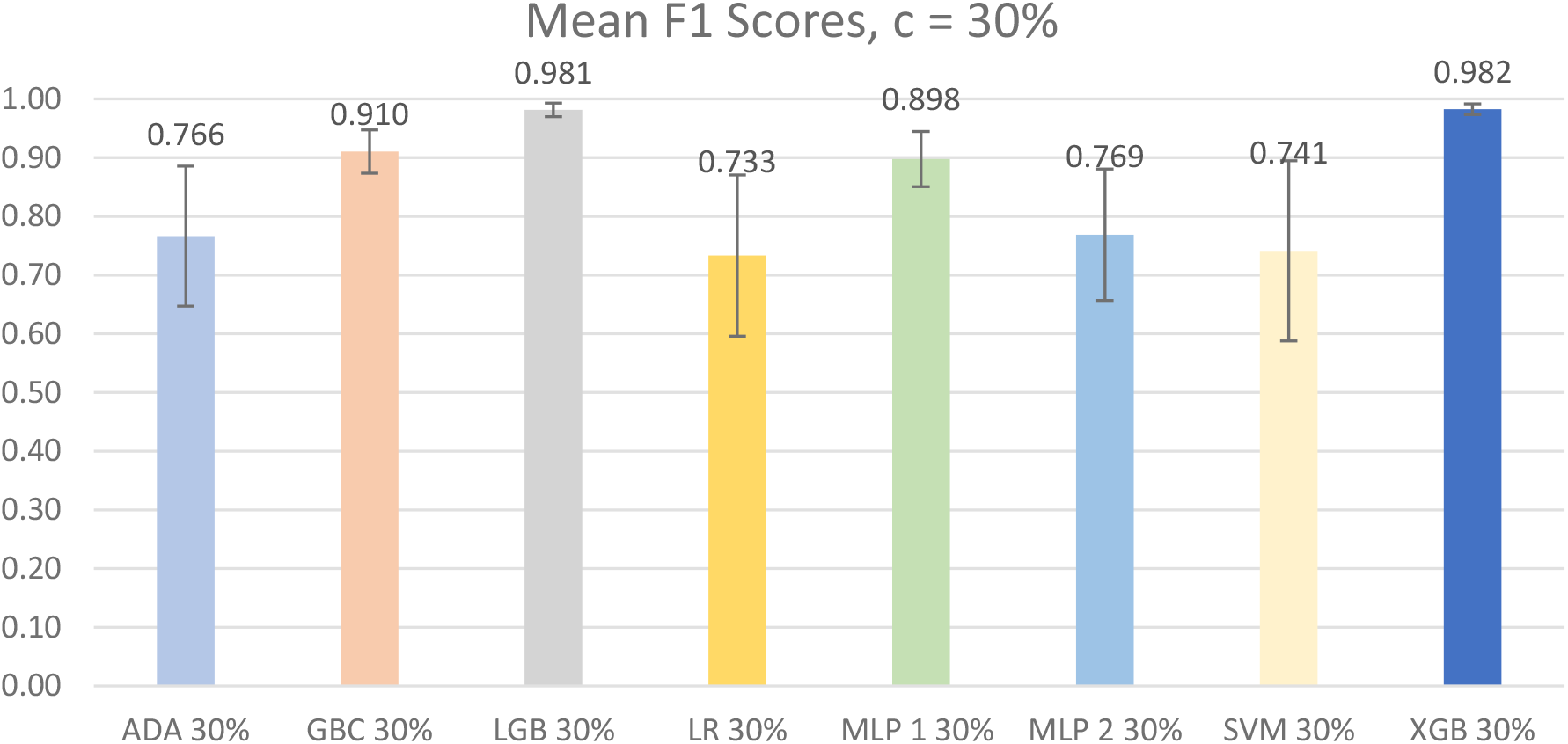
Graph of Mean F1 metrics of Left, Right, or Supine classification by model for the incremental learning level c=30%. Error bars represent the standard deviation.

#### Mean Accuracy

According to the results of the Shapiro-Wilk and the Levene tests, the data were not normally distributed (ADA: W = 0.827, p = 0.415) and were not homogenous F(7, 64) = 3.596, p < 0.01. Therefore, significant differences between the mean accuracies of the different models were found using Friedman’s ANOVA (χ^2^(7) = 54.366, p < 1×10^−8^).

Post-hoc tests with Bonferroni corrections for multiple comparisons were used to compare the performance of the top three models (GBC, LGB, and XGB). In total, three comparisons were made, changing the p-value needed to reach significance to p<0.017. A Wilcoxon Signed-Rank test was used for all three post-hoc comparisons and two were found to be statistically significant, GBC vs LGB: V = 0, p < 0.01; XGB vs GBC: V = 0, p < 0.01; LGB vs XGB: V = 13, p = 0.933. These results suggest that LGB and XGB were the two best performing models as they were both significantly different from GBC and that they should both be considered moving forward as there are no significant differences in performance between the two.

#### F1 Scores

According to the results of the Shapiro-Wilk and the Levene tests, the data were normally distributed and were not homogenous F(7, 64) = 3.682, p < 0.01. Significant differences between the mean accuracies of the different models were found using a Friedman’s ANOVA (χ^2^(7) = 54.148, p < 1×10^−8^).

Post-hoc tests with Bonferroni corrections for multiple comparisons were used to compare the performance of the top three models (GBC, LGB, and XGB). In total, three comparisons were made, changing the p-value needed to reach significance to p<0.017. A Wilcoxon Signed-Rank test was used for all three post-hoc comparisons and two were found to be statistically significant, GBC vs LGB: V = 0, p < 0.01; XGB vs GBC: V = 0, p < 0.01; LGB vs XGB: V = 21, p = 0.910. These results suggest that LGB and XGB are the two best performing models as they are both significantly different from GBC and that they should both be considered moving forward as there are no significant differences in performance between the two.

### 3.3 Incremental Learning Levels

Classifier performance for all incremental learning levels (c) for each of the top two models were compared to their adjacent c values (i.e., c = 0% to 10%, c = 10% to 20%, and c = 20% to 30%).

#### Mean Accuracy

The assumption of normality was not met for either LGB or XGB. For the LGB data, the Shapiro-Wilk test was significant for one of the cases, c = 10%: W = 0.827; p = 0.0410. For the XGB data, the Shapiro-Wilk test was also significant for one of the cases, c = 10%: W = 0.802; p = 0.0216. The assumption of homogeneity was not met for both models, LGB: F(3,32) = 12.575, p <1×10^−4^; XGB: F(3,32) = 12.764, p < 1×10^−4^. A Friedman’s ANOVA was used to determine if there was a significant difference between the means as the data did not satisfy the assumptions for a parametric test. The test reported significant differences for both the LGB and XGB models, LGB: (χ^2^(3) = 27, p < 1×10^−5^); XGB: (χ^2^(3) = 27, p < 1×10^−5^). Figure 5 shows a visual comparison between the different incremental learning levels for the LGB and XGB models.

**Figure 5.**
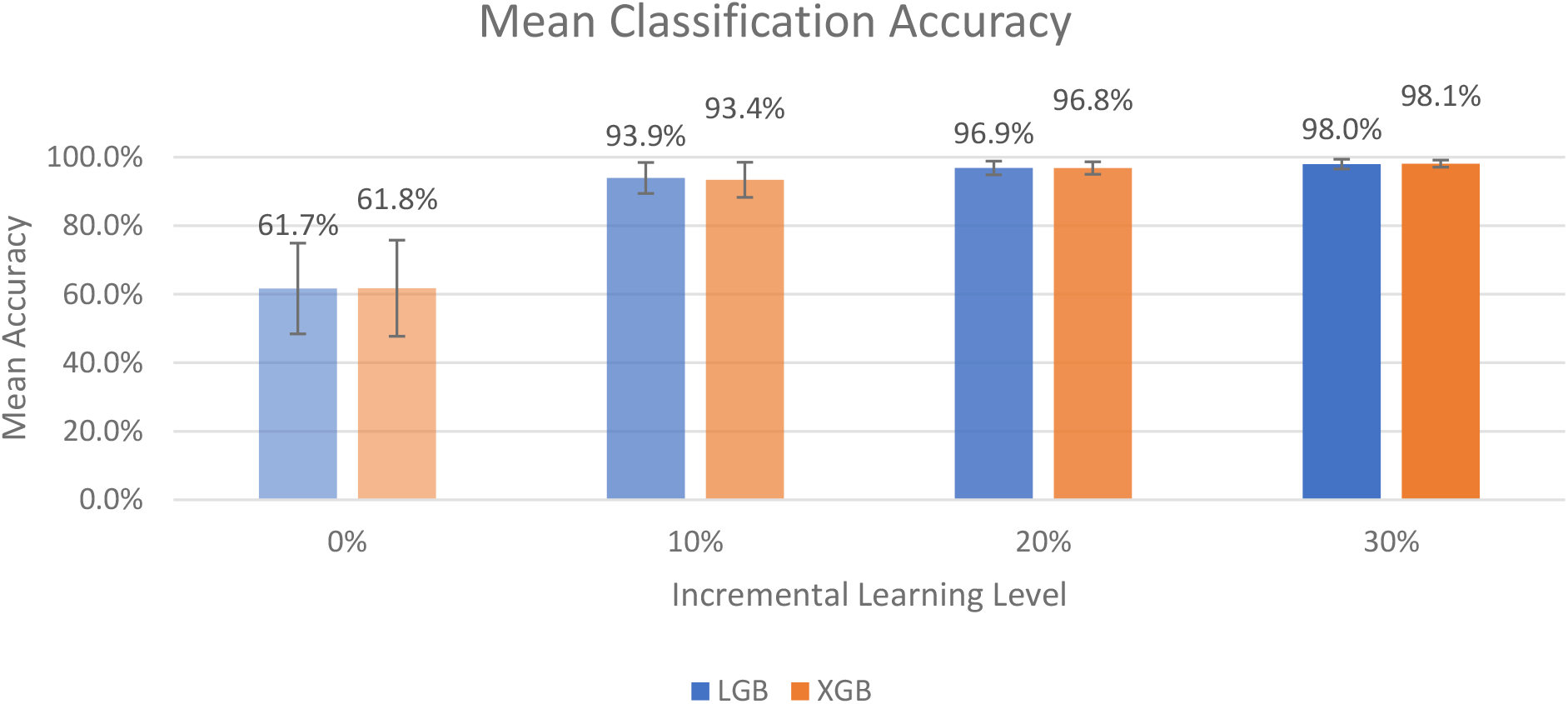
Graph of Left, Right, or Supine classification mean accuracy for the LGB and XGB models across the four different incremental learning levels. The error bars represent standard deviation.

Post-hoc tests with Bonferroni corrections for multiple comparisons were used to compare the performance of the models based on the incremental learning level used (c = 0%, 10%, 20%, or 30%). In total, three comparisons were made between adjacent incremental learning levels, changing the p-value needed to reach significance to p<0.017. A Wilcoxon Signed-Rank test was use for all three post-hoc comparisons for each model and all three comparisons were significant. For the LGB, 0% vs 10%: V = 0, p < 0.01; 10% vs 20%: V = 0, p < 0.01; 20% vs 30%: V = 0, p < 0.01. For the XGB, 0% vs 10%: V = 0, p < 0.01; 10% vs 20%: V = 0, p < 0.01; 20% vs 30%: V = 1, p < 0.01. These results suggest that for both LGB and XGB, there are significant differences in performance between each incremental learning level.

#### Mean F1 Scores

The assumption of normality was met for both LGB or XGB. The assumption of homogeneity was not met for either of the models, LGB: F(3,32) = 10.079, p <1×10^−4^; XGB: F(3,32) = 7.962, p < 1×10^−3^. A Friedman’s ANOVA was used to determine if there was a significant difference between the means as the data did not satisfy the assumptions for a parametric test. The test reported significant differences for both the LGB and XGB models, LGB: (χ^2^(3) = 27, p < 1×10^−5^); XGB: (χ^2^(3) = 27, p < 1×10^−5^). Figure 6 shows a visual comparison between the different incremental learning levels for the LGB and XGB models.

**Figure 6.**
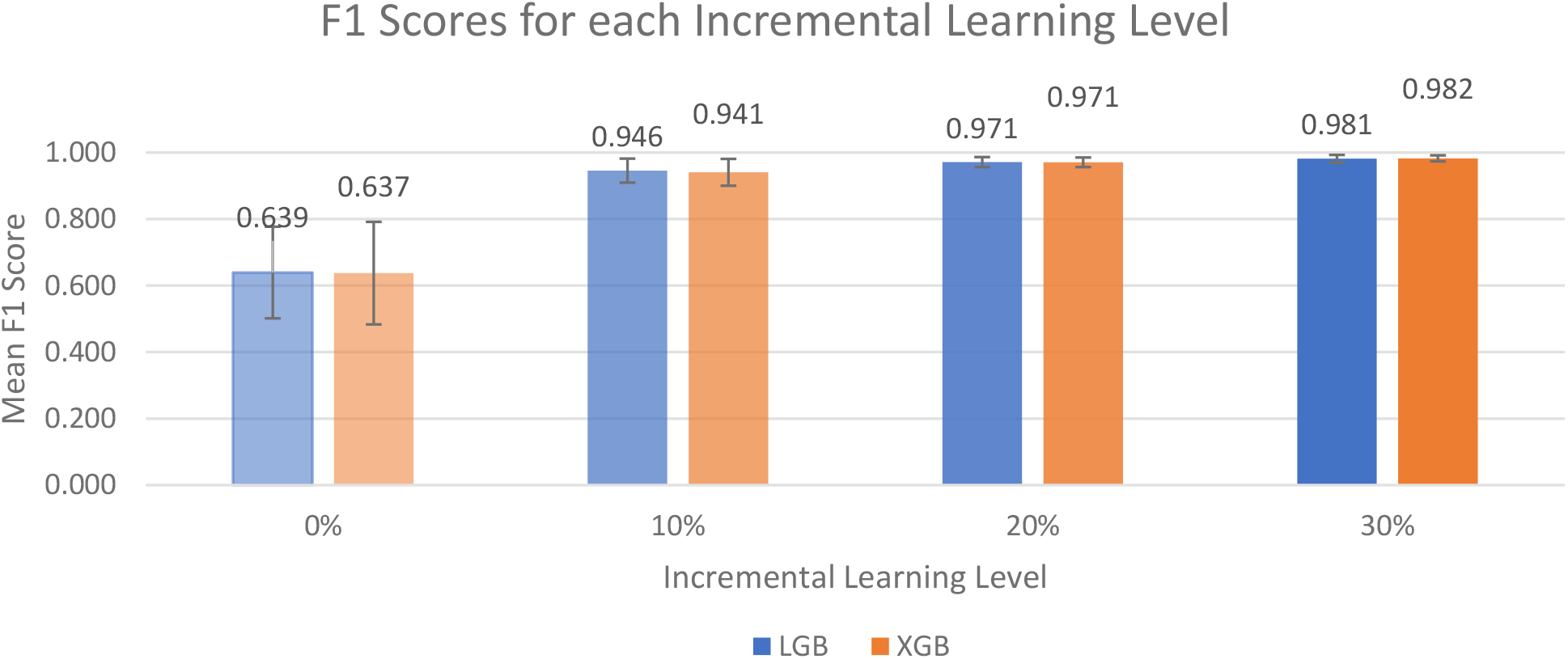
Graph of Left, Right, or Supine classification mean F1 metrics for the LGB and XGB models across the four different incremental learning levels. The error bars represent standard deviation.

Post-hoc tests with Bonferroni corrections for multiple comparisons were used to compare the performance of the models based on the incremental learning level used (c = 0%, 10%, 20%, or 30%). In total, three comparisons were made between adjacent incremental learning levels, changing the p-value needed to reach significance to p<0.017. A Wilcoxon Signed-Rank test was use for all three post-hoc comparisons for each model and all three comparisons were significant. For the LGB, 0% vs 10%: V = 0, p < 0.01; 10% vs 20%: V = 0, p < 0.01; 20% vs 30%: V = 0, p < 0.01. For the XGB, 0% vs 10%: V = 0, p < 0.01; 10% vs 20%: V = 0, p < 0.01; 20% vs 30%: V = 1, p < 0.01. These results suggest that for both LGB and XGB, there are significant differences in performance between each incremental learning level.

### 3.4 Inter-rater Reliability

In total, 13,750 images were used for reliability estimates of labelling ground truth data. Results of the inter-rater reliability analysis can be found in Table 7. Kappa values for individual positions and for each participant are provided along with the number of data points included in the analysis.

**Table 7.** Results of the inter-rater reliability analysis done using Fleiss’ kappa (κ)

| Participant | Supine ($\kappa$) | Right-side ($\kappa$) | Left-side ($\kappa$) | Combined ($\kappa$) | n |
| --- | --- | --- | --- | --- | --- |
| 1 | 0.997 | 0.988 | 0.998 | 0.991 | 1754 |
| 2 | 0.984 | 0.998 | 0.990 | 0.988 | 1684 |
| 3 | 0.970 | 0.975 | - | 0.945 | 1435 |
| 4 | 0.836 | 0.961 | 0.997 | 0.934 | 996 |
| 5 | 0.984 | 0.996 | 1.000 | 0.994 | 1419 |
| 6 | 0.989 | 1.000 | 0.999 | 0.995 | 1944 |
| 7 | 0.769 | 0.974 | 0.999 | 0.959 | 820 |
| 8 | 0.859 | 0.861 | 1.000 | 0.880 | 1295 |
| 9 | 1.000 | 1.000 | 1.000 | 1.000 | 737 |
| **Overall ($\kappa$)** | **0.969** | **0.948** | **0.960** | **0.935** | **13750** |

## 4 DISCUSSION

This study is the first that we are aware of that has explored detecting an individual’s position while they slept in the own beds at home. This study was conducted on healthy participants to determine the feasibility of this methodology prior to testing individuals at high risk of developing pressure injuries in their homes. We used ground truth comparisons to determine the participant’s true position in bed while assessing the reliability of ground truth comparisons between three different raters.

### 4.1 Machine Learning Models

In our first comparison looking at all the models, we concluded that the two best performing models were LGB and XGB at 30% incremental learning, where we achieved mean accuracies of 98.0% ± 1.42% and 98.1% ± 1.03%, respectively, and mean F1 scores of 0.981 ± 0.0116 and 0.982 ± 0.0030, respectively. This is an improvement from our previous study by Wong et al., that achieved 94.2% accuracy in a controlled lab setting, and an improvement from the ∼88.58% accuracy and 0.898 F1 score that the model from Wong et al., performed on the current home dataset. We need to establish a larger sample size and recruit more representable patients in order to further assess which model performs best and to improve generalizability.

When comparing all the models for the 30% incremental learning level, the Friedman’s ANOVA was significant for both mean accuracy and mean F1 score, demonstrating a significant difference between the performance of the models. Post-hoc comparisons using Wilcoxon Ranked-Sign tests with Bonferroni corrections were performed to compare the top three models for a total of 3 comparison: XGB vs GBC, LGB vs GBC, and XGB vs LGB. Results showed that there were statistically significant differences in mean accuracy and mean F1 score for XGB vs GBC and LGB vs GBC cases, and no statistically significant difference in the XGB vs LGB case. Therefore, the XGB and LGB models perform significantly better than all our other models and perform comparably to each other, suggesting these are the best models that are best to use in the future.

### 4.2 Incremental Learning

When comparing the incremental learning levels between the top two models, LGB and XGB, a Friedman’s ANOVA showed significant differences for both models. Post-hoc comparisons with a Wilcoxon Signed-Rank test with Bonferroni corrections were used to compare adjacent incremental learning values (i.e., 0% to 10%, 10% to 20%, and 20% to 30%) for a total of three comparisons per model. For both the LGB and XGB models, all three comparisons were statistically significant for mean accuracy and mean F1 score, suggesting that the best performing models were at an incremental learning level of 30%. Considering the biggest improvements in performance occurred between the 0% to 10% incremental learning levels (∼61% to ∼93% for mean accuracy and ∼0.63 to 0.94∼ for mean F1 score) and improvements between other levels were much smaller (<4% and 0.04). However, it is unclear if these differences are clinically significant. It may be desirable to select a model with slightly lower accuracy if it also requires less data to be collected for incremental learning with each new end user.

### 4.3 Ground Truth Labelling

The challenge of detecting an individual’s position in bed becomes difficult to detect if the individual’s body is occluded by covers (e.g. sheets, blankets) [9]. However, the results of this study demonstrated that our raters were able to detect the individual’s position by visual inspection consistently (Kappa 0.859-1). These findings demonstrate that a single rater is sufficient for providing ground truth labels in future work.

### 4.4 Future Work

Our ultimate goal of this work is to use our non-contact sensor system as an objective and unobtrusive tool for tracking repositioning in the home environment to gain a better understanding of how often patients are repositioned and to evaluate interventions that may improve adherence to regular repositioning. These interventions may include combinations of education, prompting systems, pain management, as well as paid caregiver support in the home. Our work may include extending the functionality of our sensor system to create a repositioning prompting system, designed to provide prompts to a caregiver when the patient has remained in the same position for too long.

## 5 CONCLUSIONS

The results of this study found that our system was able to detect the position of nine participants at home in their own beds (as either right-side lying, left-side lying or supine) with 98.1% accuracy and an F1 score of 0.982 using the XGBoost classifier. An inter-rater reliability analysis done using Fleiss’ kappa found almost perfect agreement (κ = 0.935) between the ground truth position labels created by our three independent reviewers.

These results demonstrate our system has excellent potential for use in the home environment for tracking patient position continuously for pressure injury prevention and management.

## Data Availability

All data produced in the present study are available upon reasonable request to the authors

## Acknowledgements

The authors would like to thank Gordon Wong, Hamed Ghomashchi, Zeyad Ghulam and Haashim Shahzada for their help with this work. Nikola Pupic received funding from the Canadian Institutes for Health Research and Sharon Gabison was awarded the Hallisey Family Affiliate Scientist Position in Pressure Injury and Home Care Research.

